# The central response of electroacupuncture for anxiety in people with obesity based on resting-state functional magnetic resonance imaging: a protocol for a randomized, blinded, sham-controlled trial

**DOI:** 10.1101/2024.09.19.24313948

**Authors:** Qi Shu, Qiumei Peng, Wenxiu Duan, Fan Zhang, Qing Yu, Ronglin Cai

**Author notes:** Qi Shu and Qiumei Peng contributed equally to this work.

## Abstract

**Introduction:** Obesity is a problem that is increasing worldwide and anxiety is a major psychological disorder associated with obesity. Electroacupuncture (EA) has been proved to be a feasible treatment for obesity and anxiety respectively in clinical practice. However, to date, there are no data on EA’s effectiveness on anxiety for people with obesity. Therefore, this study aims to evaluate the efficacy and safety of EA for anxiety in people with obesity, and to observe the brain functional status of patients and the intervention effects of EA on brain function by functional magnetic resonance imaging (fMRI).

**Methods and analysis:** This study is a randomized, blinded, sham-controlled and clinical trial. 72 obesity patients from two hospitals with anxiety will be randomly divided into EA group and control group in 1:1 ratio by using a random number table. Patients in EA group will receive EA treatment at specific acupoints with penetrating needling for 8 weeks. The control group will receive Park’s acupuncture with non-penetrating needling. Weight, waist, BMI (Body Mass Index), SAS (Self-rating Anxiety Scale), STAI (State-Trait Anxiety Inventory) and PSQI (Pittsburgh Sleep Quality Index) will be performed before, after 8-week treatments and at one-month follow-up in patients. Objective metabolic parameters such as triglyceride, total cholesterol, fasting blood glucose, ghrelin, leptin, cortisol and adrenocorticotropic hormone will also be detected before and after 8-week intervention. 20 patients will be randomly selected from EA group and control group, respectively, before treatment, and 20 paired healthy participants will be recruited at the same time. The 20 patients will be scanned by fMRI before and after treatment while the other 20 healthy participants will be scanned only at baseline. Regional homogeneity, amplitude of low-frequency fluctuation and resting-state-functional-connectivity will be carried out to compare the dysfunctional brain regions between patients and healthy participants, as well as the differences between two groups of patients after treatment.

**Ethics and dissemination:** The study protocol has been approved by the Hospital Ethics Committee of Second Affiliated Hospital of Anhui University of Chinese Medicine (2023-zj-42). Informed consent will be obtained prior to starting study- ralated procedures. The results will be disseminated in peer-reviewed journals and at scientific conferences.

**Trail Registration number:** Chinese Clinical Trail Registry. ChiCTR2400083594, registered 29 April 2024.

## Introduction

Overweight/obesity is a global issue with detrimental health impacts, which increases the incidence of type 2 diabetes, fatty liver, coronary heart disease, and anxiety or depression related mental disorders [1,2]. It is reported that more than 2.0 billion adults worldwide are overweight [3]. In China, approximately 11.1% children and adolescents aged 6-17 years are overweight and 7.9% are obese [4]. In addition, psychological factors often co-occur with obesity and may consequently maintain obesity [5,6]. For instance, high level of stress from work may change eating patterns and lead to excessive food intake toward a higher fat and sugar diet [7]. People with obesity and a disposition to anxiety, exhibit more emotional eating than normal weight people in an experimental setting. Similarly, the stigmatization of body weight and social discrimination may aggravate the anxiety and depression of obese individuals [8]. Anti-obesity medications or bariatric surgery may assist people in reaching and sustaining sufficient weight loss to meet treatment goal, but the accompanying adverse gastrointestinal effects and adverse neuropsychiatric effects such as irritability and insomnia add suffering to their lives. Although the relationship between obesity and psychological disorders is complex, studies have shown that anxiety is a common comorbidity associated with eating and weight-related disorders [9,10]. Therefore, psychological intervention has also been introduced in the multimodal management of patients with obesity.

### Rationale

Overweight/obesity constitutes a global health priority because of its prevalence and its association with numerous complications, among which anxiety is an undeniable one. People with obesity report more emotional eating than normal-weight individuals, and especially more emotional eating that is triggered by negative emotions, including anxiety and depression [11,12]. In addition, individuals who are exposed to stress for a long time may activate the hypothalamic-pituitary-adrenal (HPA) axis, leading to binge eating and a preference for high sugar and high-fat foods, resulting in overweight/obesity [13–15]. Therefore, management of overweight/obesity requires multidisciplinary treatment and a long-term care, making it a tough work.

Social demand for an effective and safe treatment of obesity, including complementary and alternative medicine has increased, and acupuncture therapy may be a promising and acceptable treatment for overweight/obesity [16]. Currently, accumulating evidences have been shown in the clinical studies and meta-analyses in terms of the effectiveness and safety of electroacupuncture treatment for weight loss [17–20]. Researchers have identified that acupuncture can adjust various metabolic functions [21], improve the fat decomposition and suppress food addiction to achieve weight loss [22]. Specially, lifestyle modification plus acupuncture treatment is more effective than lifestyle modification alone [23]. According to previous research, acupuncture combined with diet or exercise may be the most effective intervention for weight loss [24]. Diet diary, a mild diet intervention (non low-carbon diet), is used to improve the compliance of participants, and also facilitates our observation so as to eliminate the interference of diet on EA treatment. Furthermore, other evidence suggests that acupuncture exerts effect on the neuroendocrine system, modulates the immune system reducing chronic low-grade inflammation and regulates the microbiota disbalance associated with obesity [25–27].

Actually, clinical studies have confirmed that acupuncture treatment for anxiety is effective and has unique advantages [28–30]. Animal experiments have also demonstrated that acupuncture can adjust emotions by regulating the microbiota-gut- brain axis, receptors in related regions and signal pathways [31,32]. Intestinal microbiota may be a common mediator connecting obesity and emotions [33,34]. Neuroimaging results show that EA treatment increases ALFF and RSFC in regions like prefrontal cortex (PFC) involved in inhibition control, motivation, sensory and emotional processing in overweight/obesity individuals [35]. Weight gain can negatively impact long-term mental health, which providing a suitable opportunity for acupuncture treatment. Nevertheless, we could not find data to confirm acupuncture’s utility for anxiety in overweight/obese people. Moreover, the brain remains a hotbed for research identifying the nuts and bolts of the molecular machinery leading to these profound changes in emotion and body weight. Thus, further explorations need to be conducted to clarify the central mechanism of anxiety with obese patients as well as the mechanism of acupuncture treatment.

## Methods and analysis

The randomized, blinded, sham- controlled trial aims to evaluate the efficacy and safety of electroacupuncture for obese patients with anxiety, including the central mechanism of electroacupuncture treatment based on fMRI.

### Study design

This study is a prospective, randomized, participant and assessor blinded, sham- controlled, two-armed, parallel-group study with a sample of 72 patients, 36 patients in each arm. 72 potential participants from 2 hospitals in Anhui province will be recruited through advertising posters from outpatient department of endocrinology, department of acupuncture. Patients may also be referred to the trial by their doctors (general practitioners, physiotherapists or acupuncturists) and randomly divided into EA group and control group in 1:1 ratio by using a computer-generated random number table, respectively, 20 patients will be selected from each group before treatment. 20 paired healthy participants will be recruited at the same time. The results of brain fMRI scanning will be compared to trace the location of specific dysfunctional brain regions. Data for the assessment will be collected at baseline, 8- week treatment, and one-month follow-up in two groups of patients, and data in healthy group will be collected only at baseline.

This study protocol is reported in line with the Standard Protocol Items Recommendations for Interventional Trials (SPIRIT) guidelines [36], which is supplied in Supplementary File 1. This trail follows the principles of the Declaration of Helsinki (Version Edinburgh 2000). This protocol was registered in the Chinese Clinical Trail Registry (ChiCTR2400083594) and approved by the Hospital Ethics Committee of Second Affiliated Hospital of Anhui University of Chinese Medicine (2023-zj-42). Any modification in protocol is not expected unless necessary. Any changes in the selection criteria or methodology will be discussed with the entire research team and approved by the ethics committees. The trail flowchart is summarized in Figure 1 and Table 1.

**Figure 1.**
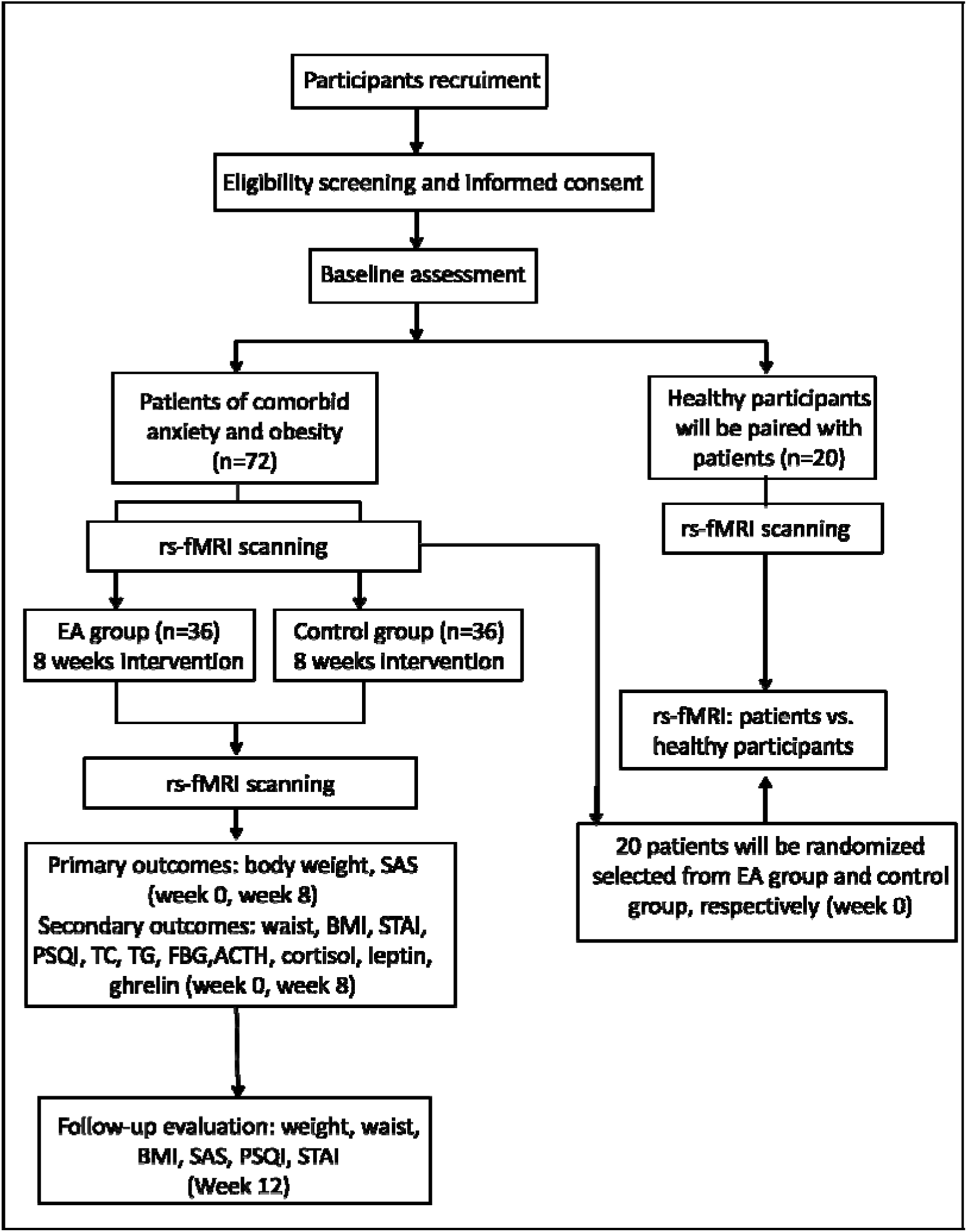
Flow diagram of study design.

**Table 1.**
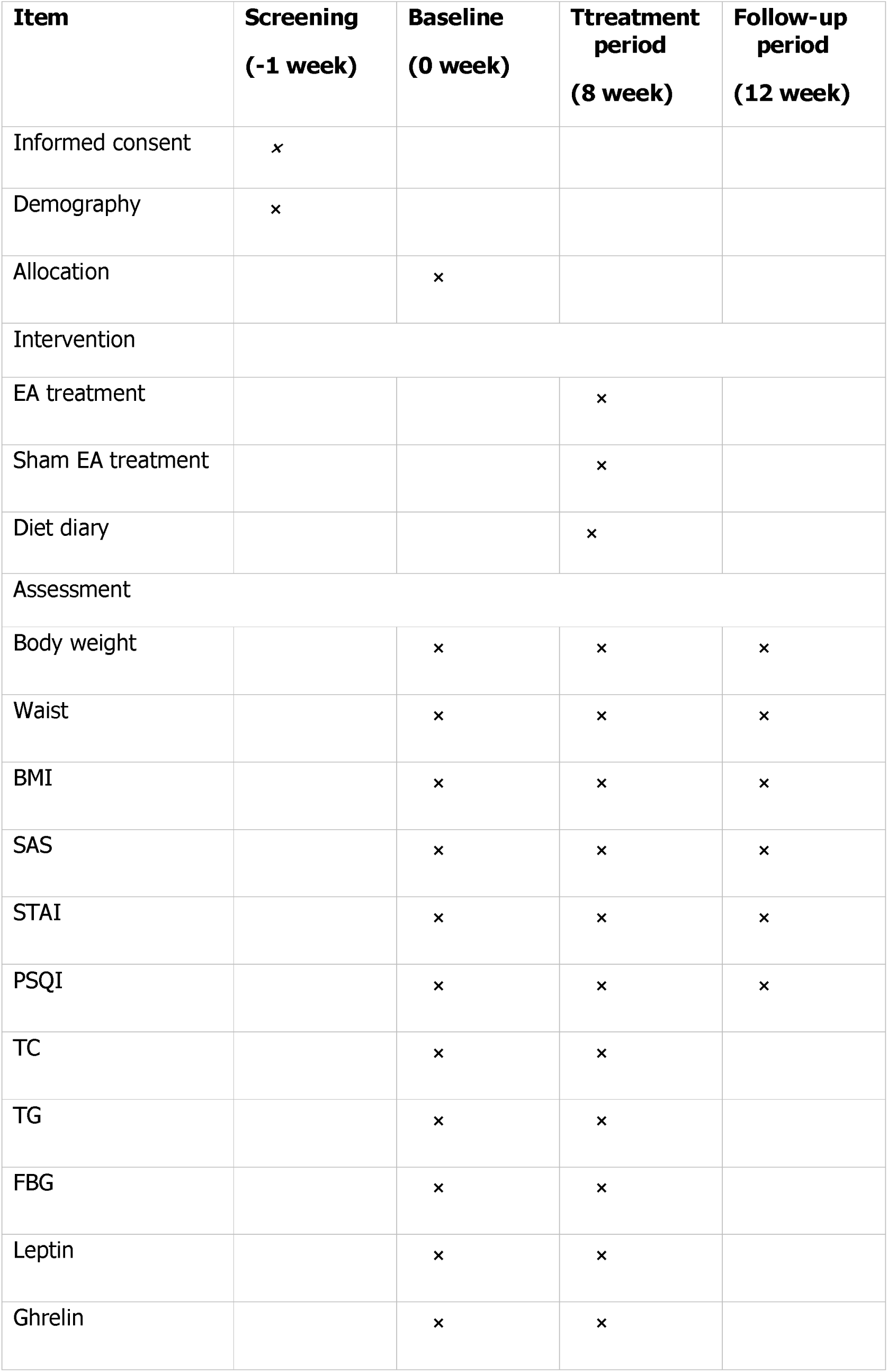
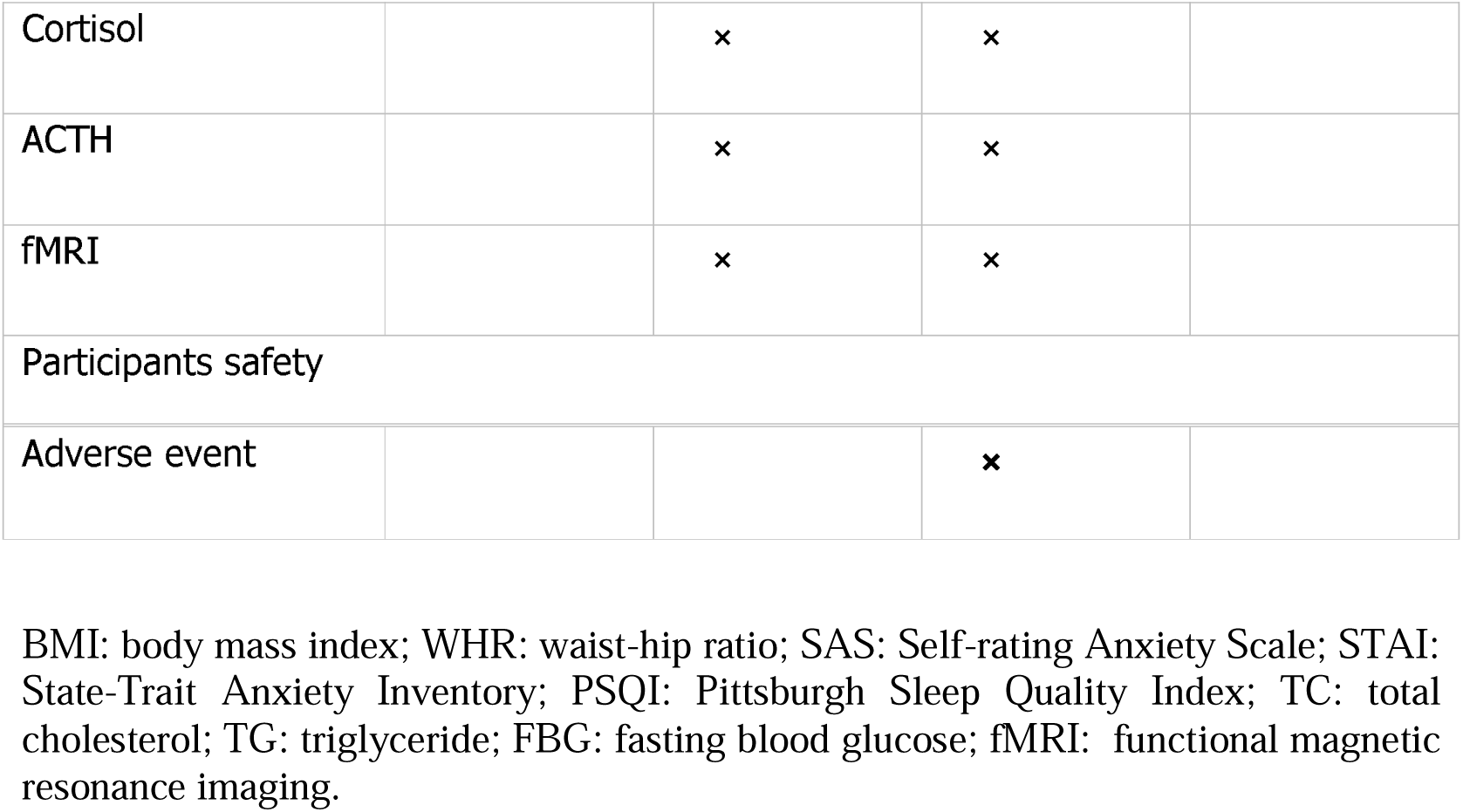
Trial Schedule.

| **Item** | **Screening**<br>**(-1 week)** | **Baseline**<br>**(0 week)** | **Ttreatment**<br>**period**<br>**(8 week)** | **Follow-up**<br>**period**<br>**(12 week)** |
| --- | --- | --- | --- | --- |
| Informed consent | x |  |  |  |
| Demography | x |  |  |  |
| Allocation |  | x |  |  |
| Intervention |  |  |  |  |
| EA treatment |  |  | x |  |
| Sham EA treatment |  |  | x |  |
| Diet diary |  |  | x |  |
| Assessment |  |  |  |  |
| Body weight |  | x | x | x |
| Waist |  | x | x | x |
| BMI |  | x | x | x |
| SAS |  | x | x | x |
| STAI |  | x | x | x |
| PSQI |  | x | x | x |
| TC |  | x | x |  |
| TG |  | x | x |  |
| FBG |  | x | x |  |
| Leptin |  | x | x |  |
| Ghrelin |  | x | x |  |
| Cortisol |  | × | × |  |
| ACTH |  | × | × |  |
| fMRI |  | × | × |  |
| Participants safety |  |  |  |  |
| Adverse event |  |  | × |  |

BMI: body mass index; WHR: waist-hip ratio; SAS: Self-rating Anxiety Scale; STAI: State-Trait Anxiety Inventory; PSQI: Pittsburgh Sleep Quality Index; TC: total cholesterol; TG: triglyceride; FBG: fasting blood glucose; fMRI: functional magnetic resonance imaging.

### Participants

This study will recruit subjects from the Second Affiliated Hospital of Anhui University of Chinese Medicine and the Anhui Province Hospital of TCM. Eligibility of patients and healthy participants will be assessed by physicians using the criteria described below and appropriately compensated after they complete the entire procedure. Informed consent will be obtained once eligible patients agree to participate in the study.

### Diagnosis criteria

This trial will adopt the diagnostic criteria of the World Health Organization for obesity in the Asia Pacific region and the diagnostic criteria for overweight/obesity in the guidelines for Prevention and Control of overweight and Obesity in Chinese Adults formulated by the Ministry of Health of China in 2003:

1. The body mass index (BMI)≥25kg/m^2^;
2. Waist circumference of men ≥85cm, waist circumference of women ≥80cm;
3. The actual measured body mass exceeds 20% of the standard body mass;

Note: After excluding secondary causes, it can be diagnosed as simple obesity if the above three criteria meet two or more.

### Inclusion criteria for patients

1. Patients who meet the diagnostic criteria for simple obesity.
2. Self-Rating Anxiety Scale (SAS) baseline score ≥ 50 points; State Trait Anxiety Inventory (STAI) baseline score ≥ 40 points.
3. Patients aged 18-45 years.
4. Patients are right-handed and regardless of gender.
5. Patients able to accept the study and sign informed consent.

### Inclusion criteria for healthy participants

20 age-matched healthy participants with similar age, sex, and educational background will be immediately recruited to pair the patients. In addition, healthy participants must meet the following criteria: physical fitness (no major illness history, no genetic history, no mental, emotional disorders, no acute or chronic disease, no cognitive impairment, no brain dysfunction or diseases, etc.), no history of fainting, no contraindication for receiving fMRI scanning, and agree to participate in the study and complete the treatment and examination as required.

### Exclusion criteria

1. Patients with endocrine system disorders and major illnesses that require treatment.
2. Pregnant or lactating women.
3. Patients who have participated in acupuncture treatment in the past month.
4. Patients who have taken weight loss, anti anxiety and depression related drugs or received weight loss treatment from a nutritionist within the past 3 months.
5. Patients with contraindications for fMRI.
6. Patients who are afraid of acupuncture and cannot cooperate with the treatment.

### Drop out criteria

1. Patients experience severe adverse reactions during the study.
2. Patients whose acupuncture treatment is interrupted for one week or more.
3. The dietary records are largely missing or incomplete.

### Randomization and blinding

Randomization will be done by SPSS (version 22.0) software in the study. The list of generated random numbers will be printed, divided into small pieces, separated and sealed in sequentially numbered envelopes. The special screeners will preserve the envelopes, and they will open the envelope when each participant is included so the group information can be gathered.

This is a patient-assessor blinded sham-controlled clinical trail. The participants will be blinded by using Prak’s non-invasive acupuncture needle (Acuprime, the United Kingdom) at the same acupoint in the same stimulation manner. Only the traditional Chinese medicine (TCM) practitioners know the allocation of each patient. The assessors and the data analyst will be blinded to the procedure and results of randomization, group allocation, and intervention. After the treatment, the participants will be asked about perceived treatment allocation to evaluate the successful rate of blinding.

### Interventions

The baseline period is one week, the treatment period is eight weeks and the follow- up is a month. Weight, waist circumference, BMI, SAS score, STAI score, PSQI score will be recorded before, after 8-week treatments and at one-month follow-up in patients. In addition, metabolic parameters such as triglyceride (TG), total cholesterol (TC), glucose will also be detected before and after 8-week intervention. The dietary records are also maintained during EA intervention. The participants in different groups will be treated separately to avoid contamination during the process. fMRI will be performed in two groups of patients at baseline (week 0), eight weeks after the treatment. Healthy participants only receive fMRI scanning at baseline. After the follow-up period, the control group will receive an eight-weeks EA treatment as compensation.

#### EA group

Acupuncture will be performed by acupuncturists with over 5 years of clinical acupuncture experience and obtained a license from the Ministry of Health of the People’s Republic of China. Before performing the treatment, acupuncturists will receive centralized training courses which include an introduction to the basic clinical research methods and a practical demonstration of the treatment. Patients will receive electroacupuncture treatment three times a week, a total of 24 times for 8 weeks. The needle retention time is 30 min at each time.

#### Acupuncture acupoints

The acupuncture point regimen is based on evidence-based clinical practice guidelines of traditional Chinese medicine. All acupoints selected and their locations are displayed in Figure 2 and Table 2.

**Figure 2.**
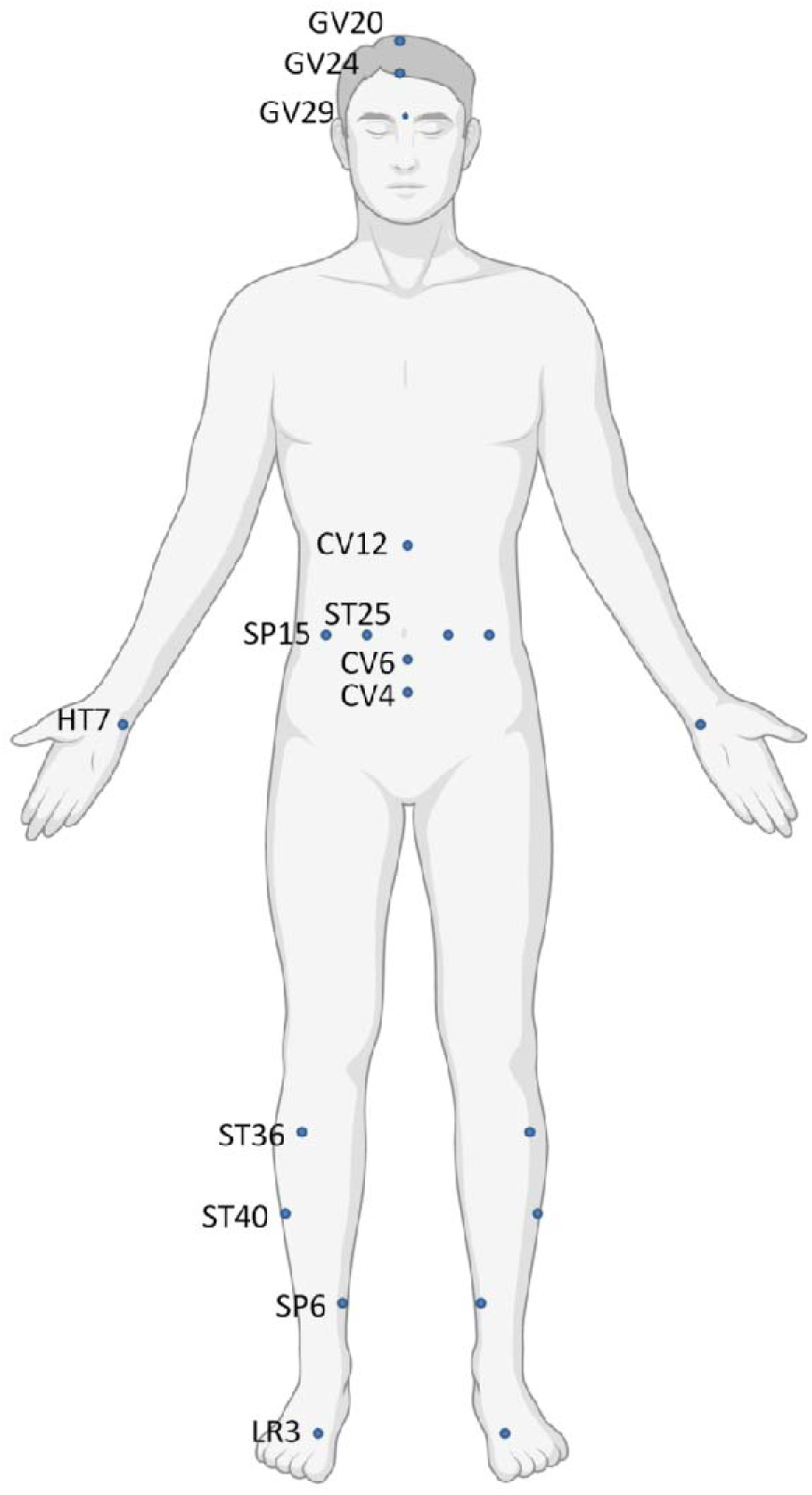
Diagram of acupoints.

**Table 2.**
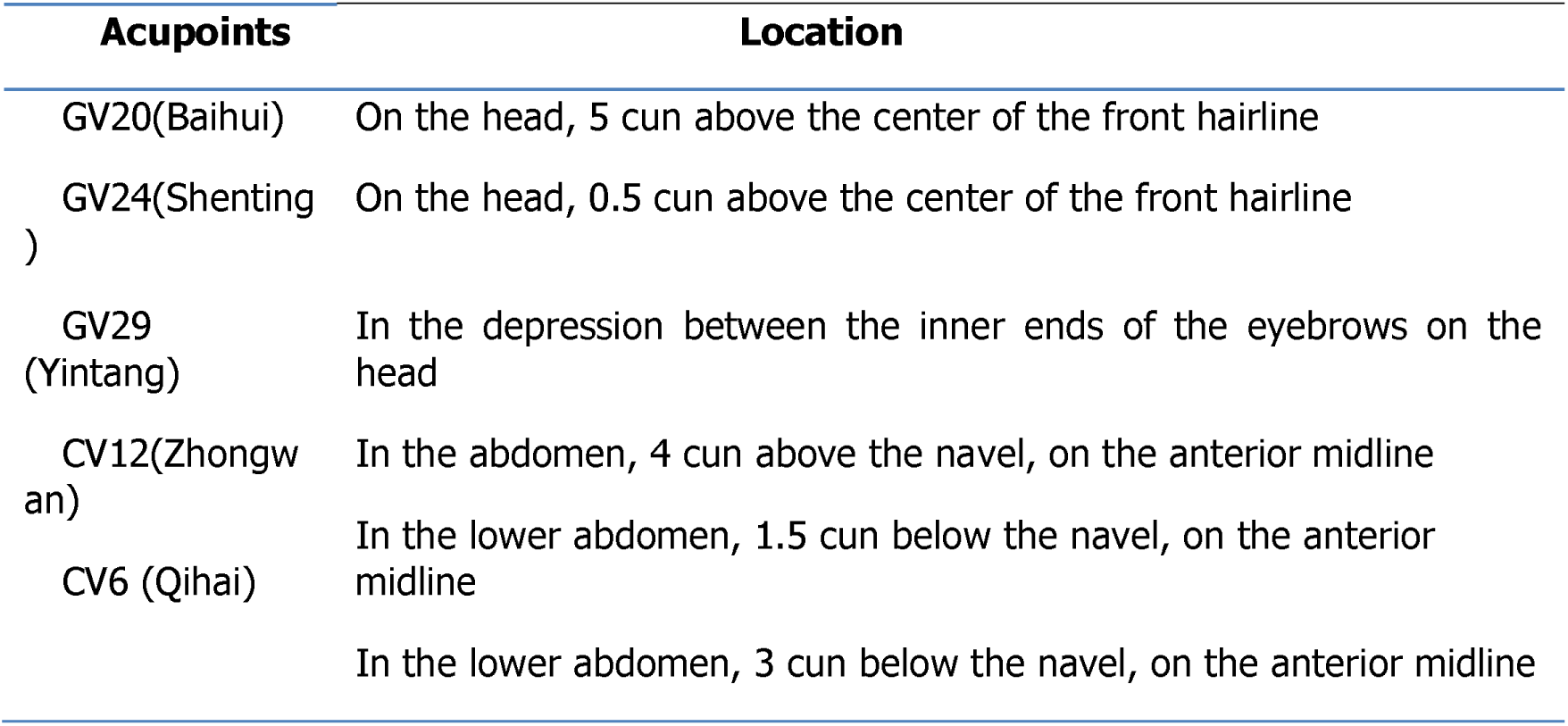
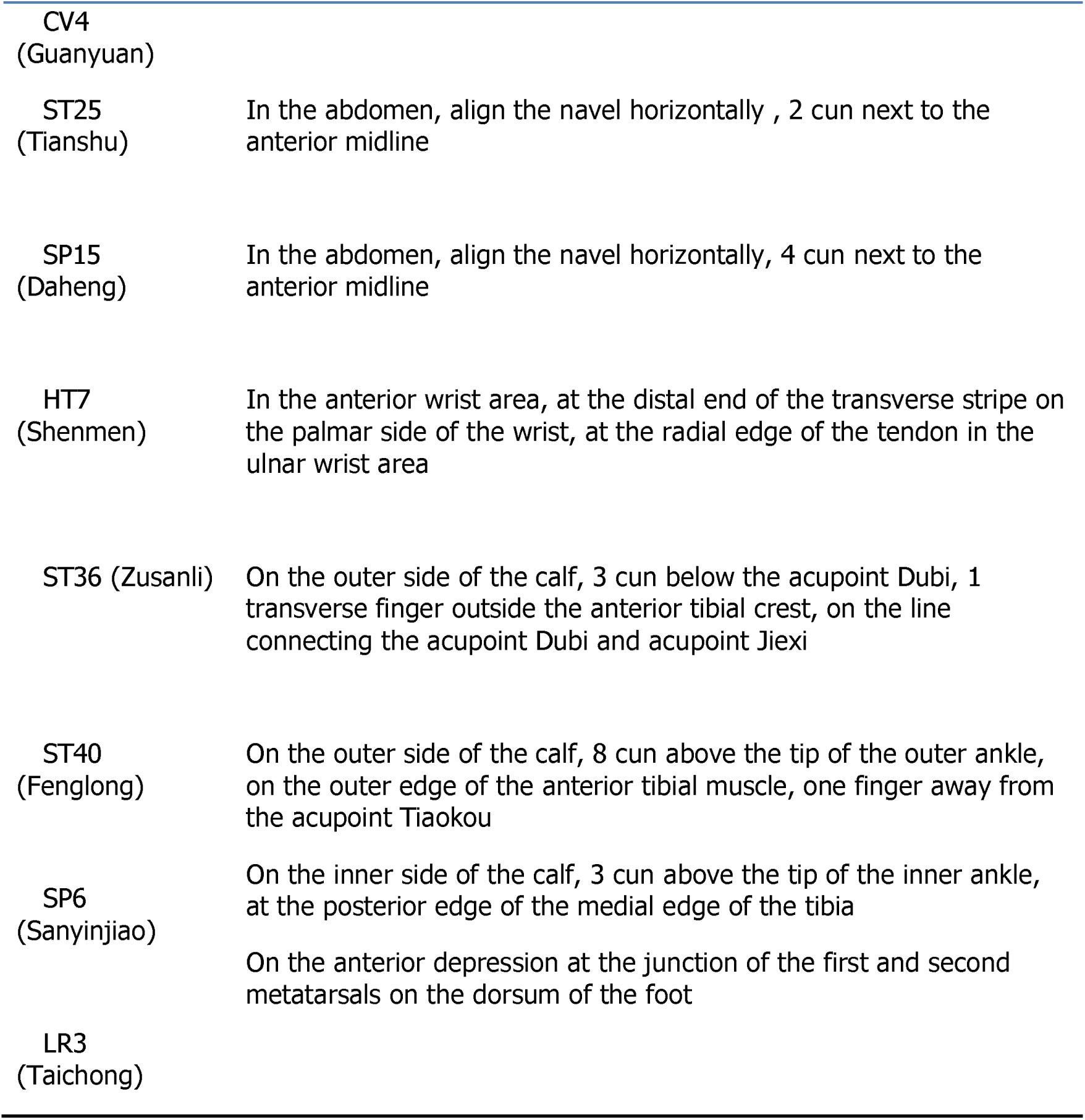
Location of acupoints.

#### Manipulation

Patients are in a supine position and the acupoints and needle are disinfected before acupuncture operation. For GV20 (Baihui), the needle will be inserted backward under the cap membrane at angle of 45° with the scalp; for GV24 (Shenting), the needle will be inserted backward under the cap membrane at an angle of 15° with the scalp; for GV29 (Yin tang), the needle will be inserted downward at an angle of 15°under the periosteum; making the patients feel heavy pressure and acid distension. For CV12 (Zhong wan), CV6 (Qihai), CV4 (Guan yuan), ST25 (Tianshu), SP15 (Daheng), HT7 (Shenmen), ST36 (Zusan i), ST40 (Feng long), SP6 (Sanyinjiao) and LR3 (Taichong), needles will be inserted at a depth of 25-35 mm vertically. The needles will be gently rotated and lifted once to achieve a sense of sourness, distention and heaviness. Electroacupuncture will then be applied to the abdominal and crural acupoints with 2/10Hz dilatational waves through electronic needle therapy device (SDZ-Ⅲ, Huatuo, China).

#### Control group

Park’s non-invasive acupuncture needle will be performed at the same acupoints in the same stimulation manner but the needles will only adhere to the skin and nit be inserted. The validity and credibility of the model have been fully proven. The acupoints in the control group will also be connected to the same electronic needle therapy device without electric current.

#### Healthy group

There will be no treatment specifically designed for the healthy group.

#### Lifestyle intervention

Dining according to the guidance of the Chinese Dietary Pagoda. All patients will be advised to take regular number of meals daily and not to intake any snacks. In addition, all participants will be required not to take exercise except for essential activities in their daily work in order to evaluate the sole effect of acupuncture treatment. The patient diet diary is supplied in Supplementary File 2.

### Outcome Measures

A printed or electronic case report form (CRF) with the random number for each participant will be used to record all information. Two assessors who are blind to the group allocations will collect the basic information and assess the outcomes from the baseline to the end of follow-up. Telephone interviews combined with offline visits are the main methods to use during the follow-up period.

#### Primary outcome measures

The primary outcomes will be the changes in the weight and SAS scores before and after the treatment.

##### Self-rating Anxiety Scale (SAS)

Self-rating Anxiety Scale (SAS) has been proposed as a useful tool for measuring the frequency of anxiety symptoms. The questionnaire adopts a four-level rating system with 20 items. Add up the scores of 20 items to obtain a rough score and multiply the rough score by 1.25 to obtain the standard score. A standard score greater than 50 indicates the presence of anxiety.

#### Secondary outcome measures

The secondary outcomes will include the changes in waist circumference, BMI, STAI scores, PSQI score during treatment and follow-up period. Total cholesterol (TC), triglyceride (TG), fasting blood glucose (FBG), cortisol, adrenocorticotropic hormone (ACTH), leptin and ghrelin will be measured before and after the 8-week treatment. Body weight, BMI and WHR are measured by body composition analyzer ( JAWON GAIA KIKO, Korea).

##### State-trait anxiety inventory (STAI)

The State-trait anxiety inventory (STAI) consists of 40 questions and is divided into two sub scales, evaluating anxiety as a transient state (state anxiety) and a latent trait (trait anxiety). The subscale score ranges from 20 to 80, with higher scores indicating higher levels of anxiety. [37,38]

##### Pittsburgh Sleep Quality Index (PSQI)

The 18-item PSQI questionnaire is a reliable and valid measure of sleep quality. The total score ranges from 0 to 21. PSQI = 7 as a reference threshold for adult sleep quality issues.

#### rs-fMRI examination and data analysis

A blood oxygenation level dependent gradient echo-planner imaging (BOLD GRE- EPI) sequence will be used to scan the whole brain, TR/TE=2000ms/30ms, turning angle = 90 ° , FOV= 220mm ×220mm, matrix = 64×64, slice thickness/spacing= 3.0/1.0 mm, layer number = 36, scanning time 8 min. All participants will lie flat on the scanning bed with their eyes closed, and without performing any tasks.

Image preprocessing will be conducted by Statistical Parametric Mapping (SPM12, http://www.fil.ion.ucl.ac.uk/spm) and the Resting-State fMRI Data Analysis Toolkit (REST, http://www.restfmri.net) on Matlab (MathWorks, Inc., Natick, MA, USA). Data preprocessing will be accomplished by SPM12: 1) remove the previous 5 unstable time points of the data; 2) time correction of the remaining time points; 3) spatial head motion correction; 4) all the subjects’ images will be spatially standardized, and re-sampled to a voxel size of 3 × 3 × 3 mm^2^; 5) remove linear drift; 6) removal of the covariates of head motion parameters and cerebrospinal fluid signal covariates; 7) remove the effects of high-frequency noise by band-pass filtering (0.01- 0.08 Hz) [39–43].

##### The regional homogeneity (ReHo) calculation

ReHo measuring the short-distance functional connectivity at voxel-level with Kendall’s coefficient of concordance (KCC), which was calculated from the consistency of the time series between each individual hormone and its 26 closest neighbors in the brain region. By calculating the KCC value of each given Voxel, the KCC diagram or ReHo diagram of each subject can be obtained. ReHo will be obtained by dividing the average KCC value of the whole brain into the KCC value of each voxel. Finally, we will smooth with a Gaussian kernel of 6 mm full-width at half-maximum (FWHM) to reduce noise [44–46].

##### The amplitude of low-frequency fluctuation (ALFF) calculation

The ALFF algorithm assumed that the low frequency BOLD signals in resting-state brain has its physiological signification. It uses the average of the amplitudes of all the frequencies in a certain frequency band (0.01 ∼ 0.08 Hz) to characterize the spontaneous activity of a voxel which reflects the level of the spontaneous activity of each voxel at rest on the view of energy point. The preprocessed time series will be first converted to a frequency domain with a fast Fourier transform and the power spectrum is obtained. The square root of the power spectrum is computed at each voxel and the averaged square root is obtained in the 0.01–0.08 Hz bandwidth at each voxel. For standardization, ALFF of each voxel is further divided by whole-brain mean ALFF values [47,48].

##### The functional connectivity (FC) calculation

The regions showing significant results in the ALFF are defined as regions-of-interest (ROIs). After obtaining ALFF-ROIs with significant main/interaction effects, a seed-region-based (center at coordinates of the peak value with a 6mm radius) RSFC-analysis is carried out. We will use band-pass filtering with a frequency range of 0.01–0.08 Hz for the RSFC calculation. Meantime- series of each ROI from the resting-state-scan is extracted, and then the strength of the RSFC for each voxel is estimated using a Pearson-correlation-coefficient between the average time-varying signal in the seed and voxel in the brain. Fisher transform is used to convert correlation maps into normally distributed coefficient maps. A two-way-ANOVA was implemented in SPM 12 to model the effects of group and time on seed-based-RSFC with an identical threshold as for ROI identification (pFWE < 0.05, cluster size of 50, cluster-forming threshold of p < 0.001). One-sample-t-test is used to test the significance of functional connections of each group of ROIs [49].

### Incidence of Adverse Events

EA is widely accepted as relatively safe if applied appropriately by competent practitioners. The type, time of occurrence, severity, and duration of the adverse events will be recorded in detail from week 0 to 8 and follow-up. Adverse events associated with EA might include fainting, stuck needle, bent needle, broken needle and hematoma, which will be reported to the Ethics Committee. The determination to discontinue the trial for a participant will be made by physicians.

Participants who suffer harm will be compensated.

### Sample Size

This study estimated the sample size using the sample size estimation formula based on the comparison of two sample means described in the following formula:

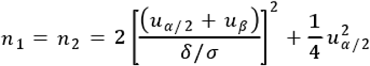. Among the formula, n_1_ and n_2_ are the required sample sizes for two groups, respectively. Based on our pre-experiment, δ/σ =0.86. In this study μ_α/2_=1.960, μ_β_= 1.282, after substituting into the above formula, each group requires 30 participants. Considering issues such as compliance and loss of follow-up during clinical trials, an additional 20% sample size was added, resulting in a total of 72 participants recruited.

### Quality Control

All staff will undergo special training on the purpose and content of the trial, treatment strategies, and quality requirements before the trail. Monitors will check the participant adverse event table, the participants’ dietary diaries and case report form once a week as well as the EA operation during the treatment period. Dropouts and withdrawals and the underlying reasons will be documented in detail throughout the trial.

### Data Collection and Management

The researchers will be required to fill the CRF with the data collected in accordance with the requirements of the study program. The researchers will submit to data management center the complete and signed CRF of all the selected patients in the study after the study is finished. The consistency of case report data collected from the study will be checked, and queries for inconsistent data will be sent to doctors for clarification. The personal CRF of each participant will be stored in a locked cabinet. All electronic documents will be stored in Anhui University of Chinese Medicine computer with password protection. Access to all research data will only be permitted to an authorized research team member.

All participants will receive a summary of study findings and group information upon trial completion. Five years after the end of the project, the electronic documents need to be deleted.

### Statistical Analysis

Based on the intention-to-treat (ITT) principle, an independent statistician will verify the effectiveness and safety of the intervention. Values missing from the dataset will be imputed using last-observation-carried-forward method.

The statistician will perform statistical analysis by using SPSS 23.0 statistics software (SPSS Inc, Chicago, IL, USA) and GraphPad 7.04 statistics software (GraphPad Software Co., San Diego, California, USA). For continuous data, the data will be presented as mean ± SD if normally distributed or as median (IQR) if not normally distributed. Longitudinal continuous data will be compared between the groups by repeated-measures analysis of variance (ANOVA). Other continuous data will be analyzed by Student’s t-test and Wilcoxon rank-sum test, and categorical data by X^2^ test or Fisher’s exact test, as appropriate. Linear correlation analysis will be conducted to measure the closeness of linear correlation between variables. *P* ≤ 0.05 will be considered statistically significant.

For ReHo analysis and ALFF analysis, 2-sample-t-tests will be conducted within the anxiety combined with obesity patients and healthy participation before treatment, as well as EA group and control group before and after treatment. Paired t-tests will be performed between two groups of patients before and after treatment to evaluate the ReHo and ALFF difference, *P* ≤ 0.05 is used to indicate a significant difference. The ALFF brain region (different brain region) to which the peak point belonged will bedetermined according to REST 1.8 software. The different brain regions of the two groups will be set as seed points, and the FC between each seed point and other brain regions are analyzed separately.

### Dissemination

The results will be published in peer-reviewed journal, a PhD thesis and presented at conferences. Data will be published in aggregate to avoid individual participant identification and presented in such a way that identifiable data are removed.

### Article Summary

#### Strengths and limitations of this study

This study may provide high-quality evidence regarding the efficacy and safety of EA for patients with obesity and anxiety comorbidities. In addition, fMRI will be introduced into clinical trials of EA intervention and provide further objective basis for the efficacy, immediate effect, cumulative effect and long-term effect of EA treatment. However, the current trail has a small sample size.

### Trial Status

Participant enrollment is started from May 2024. 40 participants have been recruited till now.

## Supporting information

https://orcid.org/my-orcid?orcid=0000-0002-7322-0798

## Abbreviations

EA: Electroacupuncture
rs-fMRI: resting-state functional magnetic resonance imaging
BMI: Body mass index
SAS: Self-rating anxiety scale
STAI: State-trait anxiety inventory
PSQI: Pittsburgh sleep quality index
TG: Triglyceride
TC: Total cholesterol
FBG: Fasting blood glucose
ReHo: Regional homogeneity
ALFF: Amplitude of low-frequency fluctuation
RSFC: Resting-state-functional-connectivity
SPIRT: Standard Protocol Items Recommendations for Interventional Trials
TCM: Traditional Chinese medicine
CRF: case report form
ITT: Intention-to-treat
HPA: Hypothalamic-pituitary-adrenal
RCT: Randomized controlled trial
PFC: prefrontal cortex.

## Data Availability

All data produced in the present study are available upon reasonable request to the authors

## Acknowledgements

We appreciate the effort and help from all of those involved in this trial.

## Author Contributions

QS is the main researcher who designed the study, drafted the manuscript and registered the trail in Chinese Clinical Trail Registry. QMP is the co-researcher who contributed to the design of the protocol and writing of the manuscript. WXD and FZ contributed to the design of the interventions. QY contributed to the statistical design and the revision of the manuscript. RLC provided the funding and revised the manuscript. All authors contributed to the revision and approved the final manuscript.

## Availability of data and materials

No data were used to support this protocol. The outcomes data of the trial will be published within the final study manuscript and will also be uploaded to the Chinese Clinical Trial Registry.

## Declarations

### Ethics approval and consent to participate

This study has been approved by the Ethics Committee of the Second Affiliated Hospital of Anhui University of Chinese Medicine (Approved number: 2023-zj-42). All participants will be informed and sign a written consent form.

### Patient and public involvement

Patients and/or the public were involved in the design, or conduct, or reporting, or dissemination plans of this research. Refer to the Methods section for further details.

### Patient consent for publication

Not applicable.

### Provenance and peer review

Not commissioned; externally peer reviewed.

## Reporting checklist for protocol of a clinical trial

Based on the SPIRIT guidelines.

### Instructions to authors

Complete this checklist by entering the page numbers from your manuscript where readers will find each of the items listed below.

Your article may not currently address all the items on the checklist. Please modify your text to include the missing information. If you are certain that an item does not apply, please write "n/a" and provide a short explanation.

Upload your completed checklist as an extra file when you submit to a journal.

In your methods section, say that you used the SPIRITreporting guidelines, and cite them as:

Chan A-W, Tetzlaff JM, Gøtzsche PC, Altman DG, Mann H, Berlin J, Dickersin K, Hróbjartsson A, Schulz KF, Parulekar WR, Krleža-Jerić K, Laupacis A, Moher D. SPIRIT 2013 Explanation and Elaboration: Guidance for protocols of clinical trials. BMJ. 2013;346:e7586

---

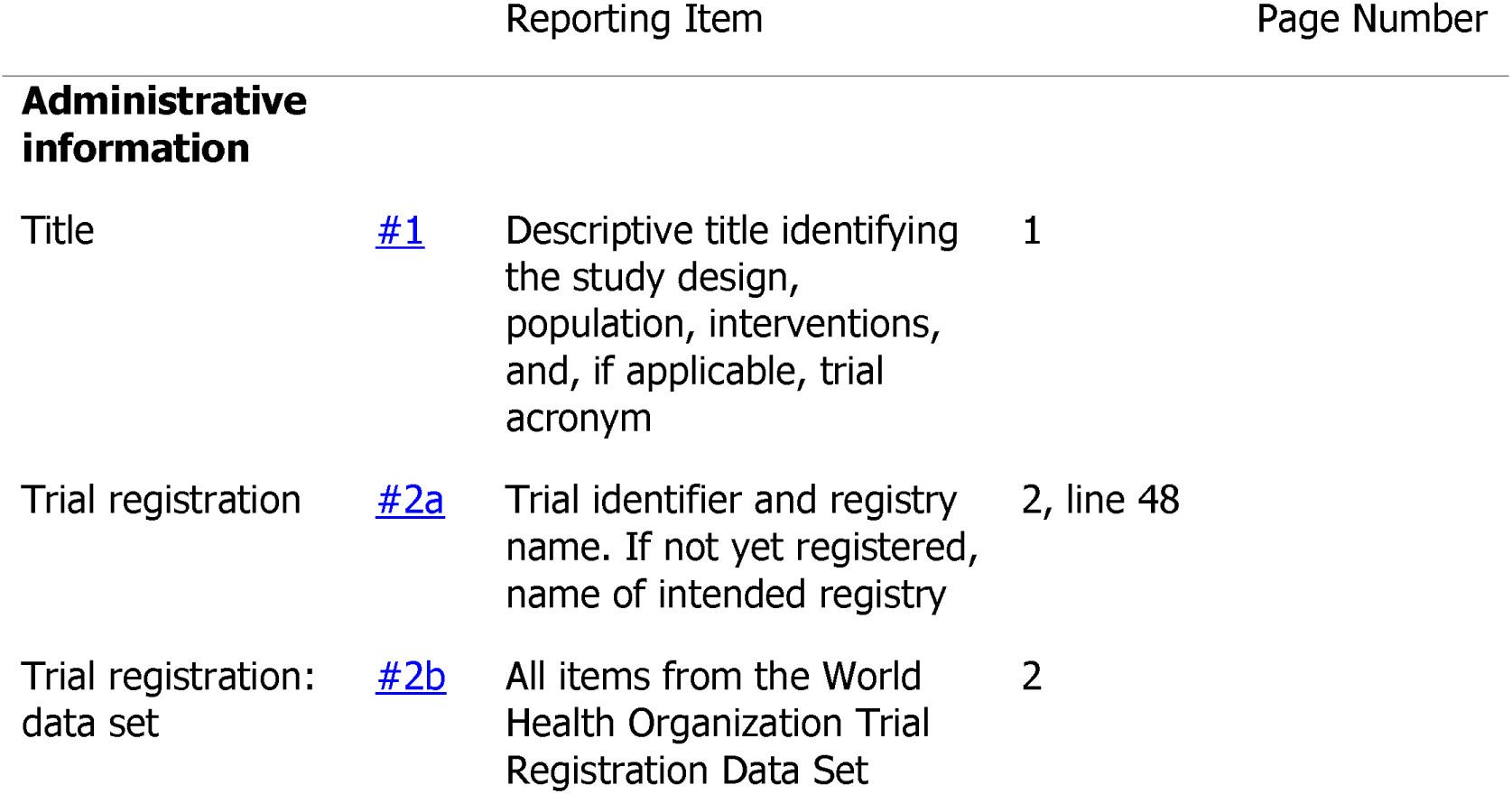
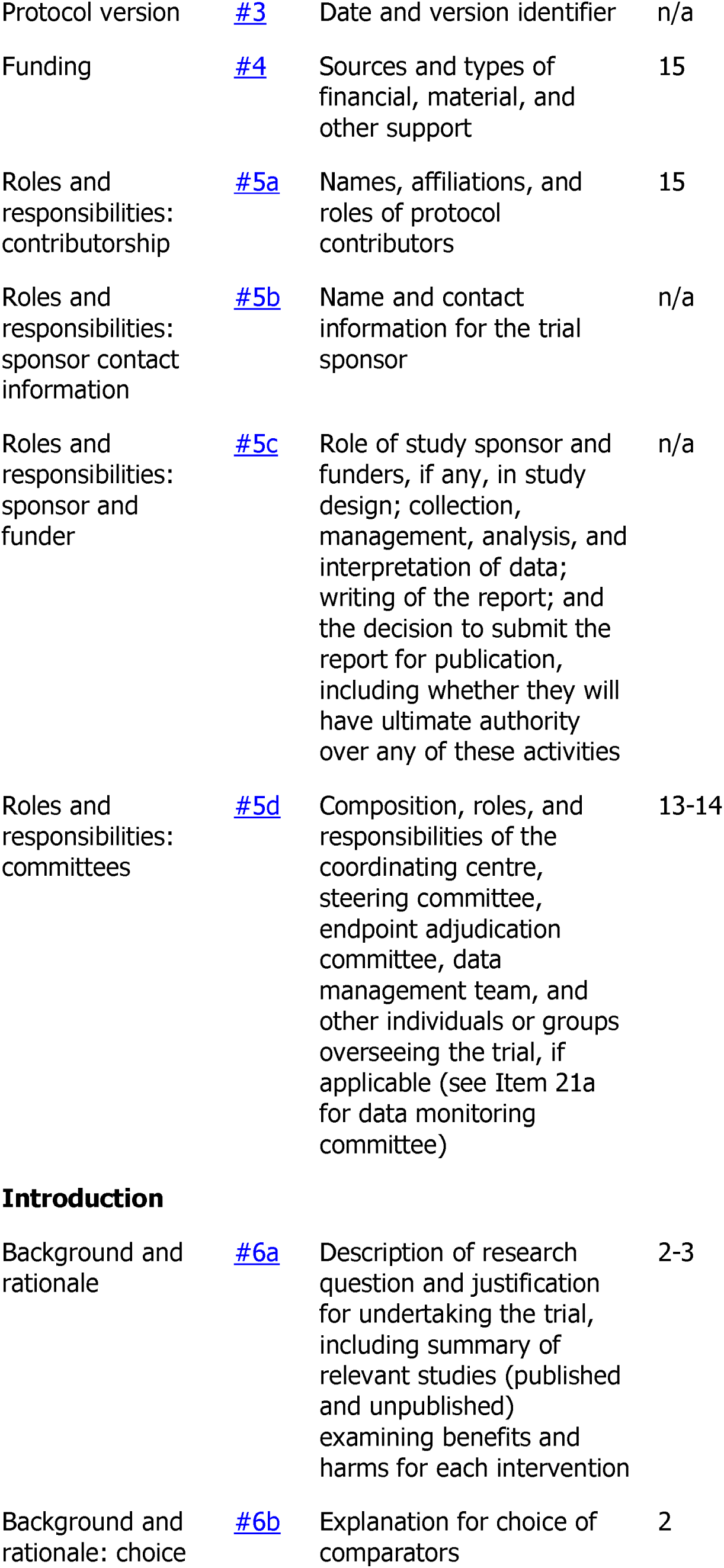
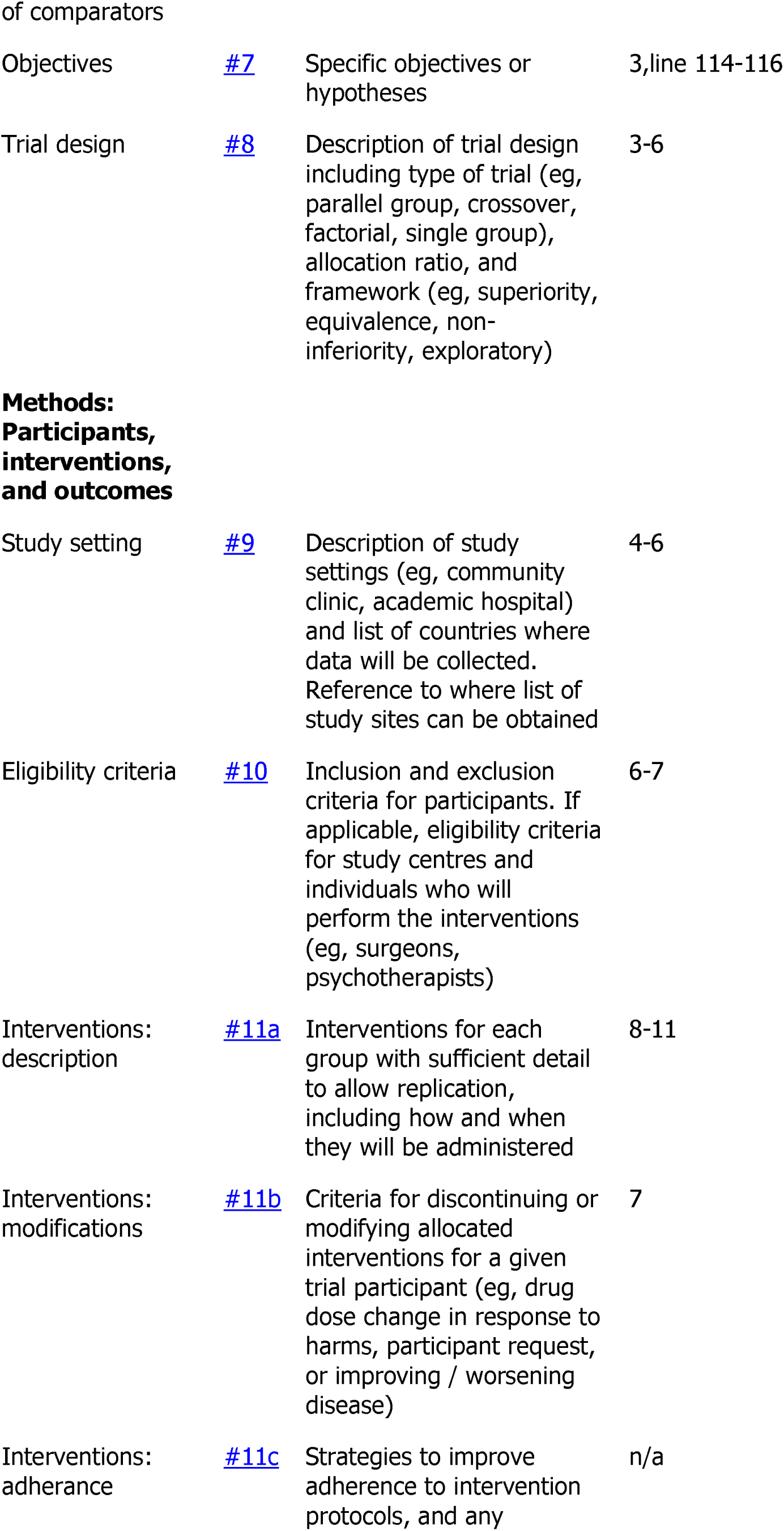
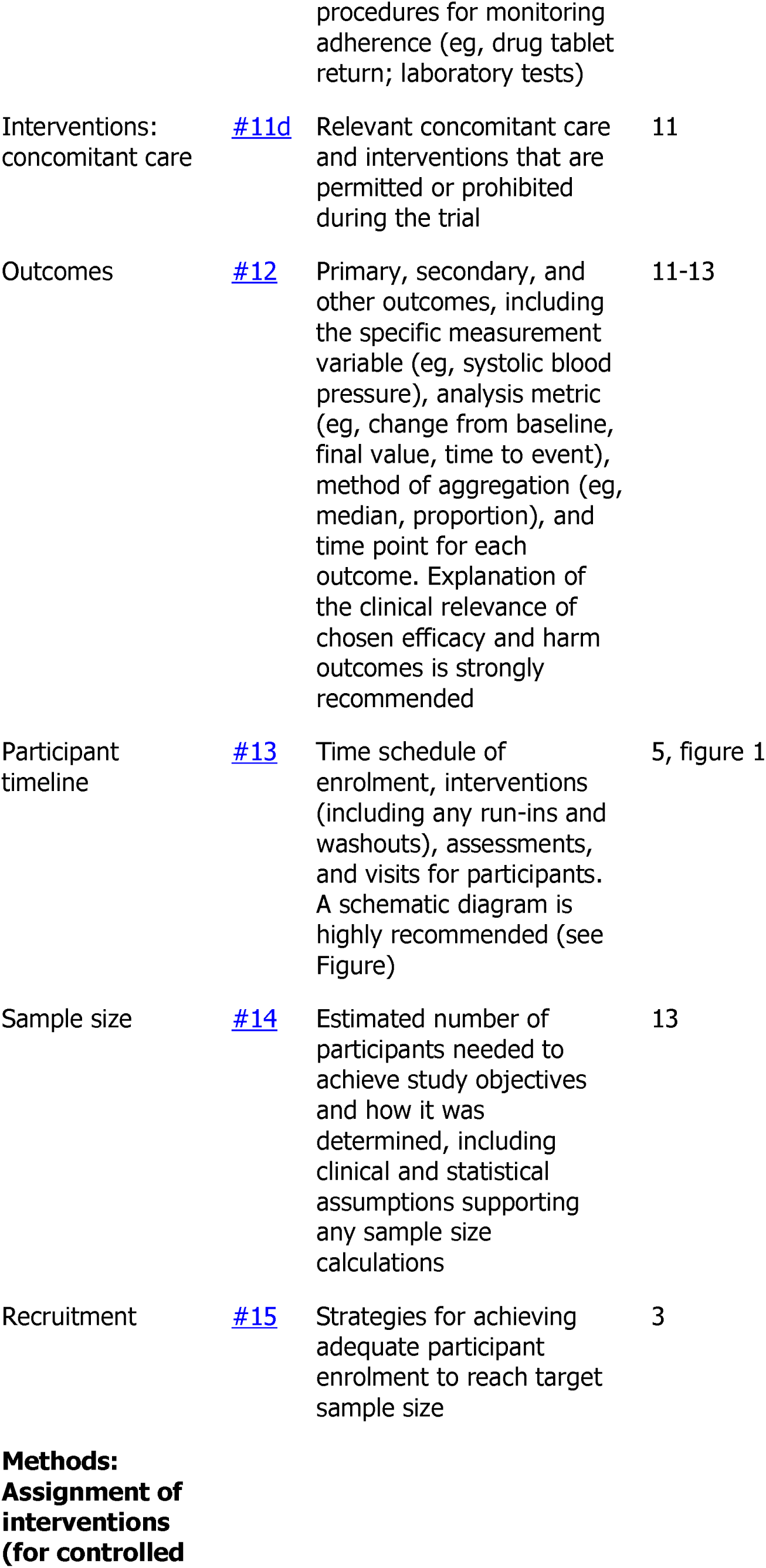
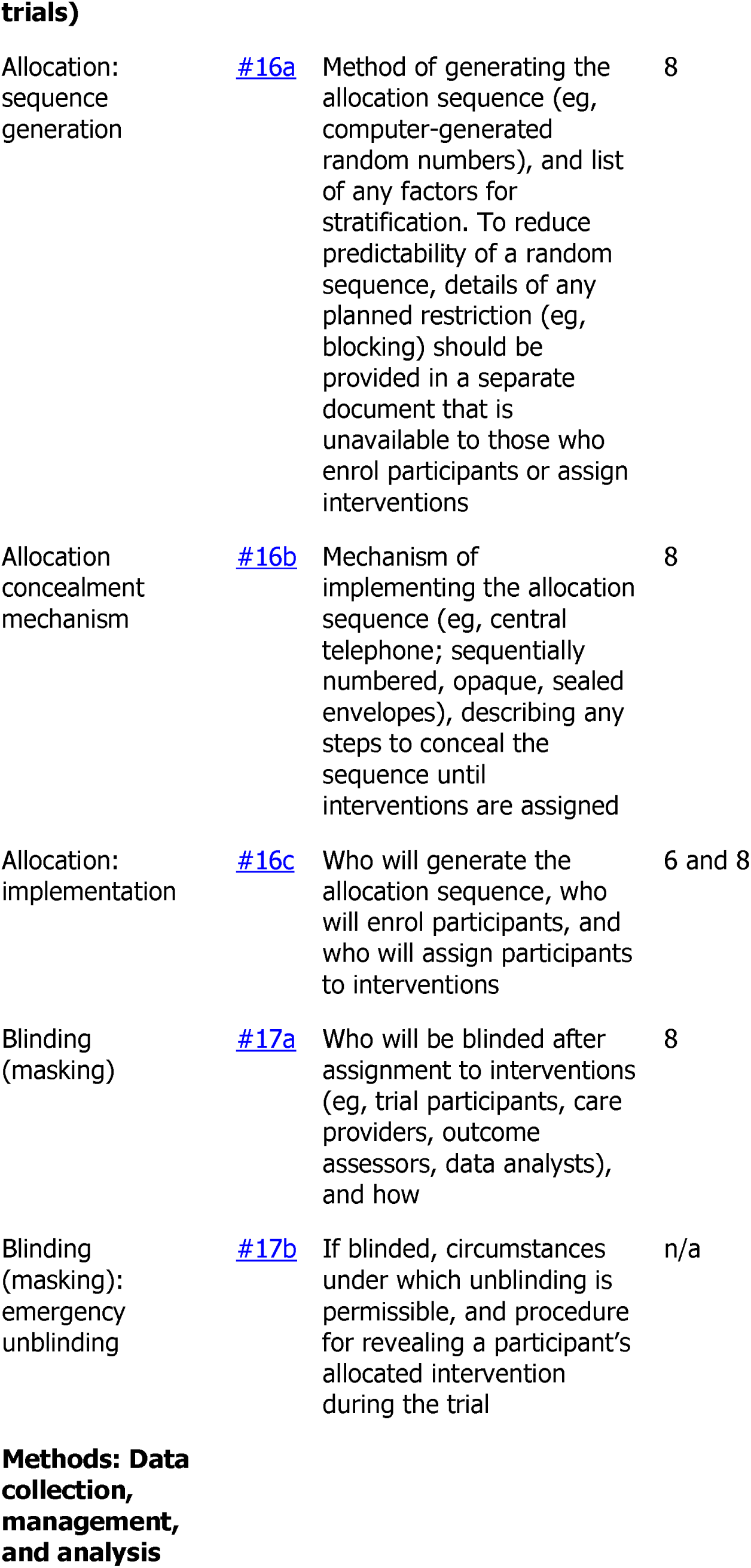
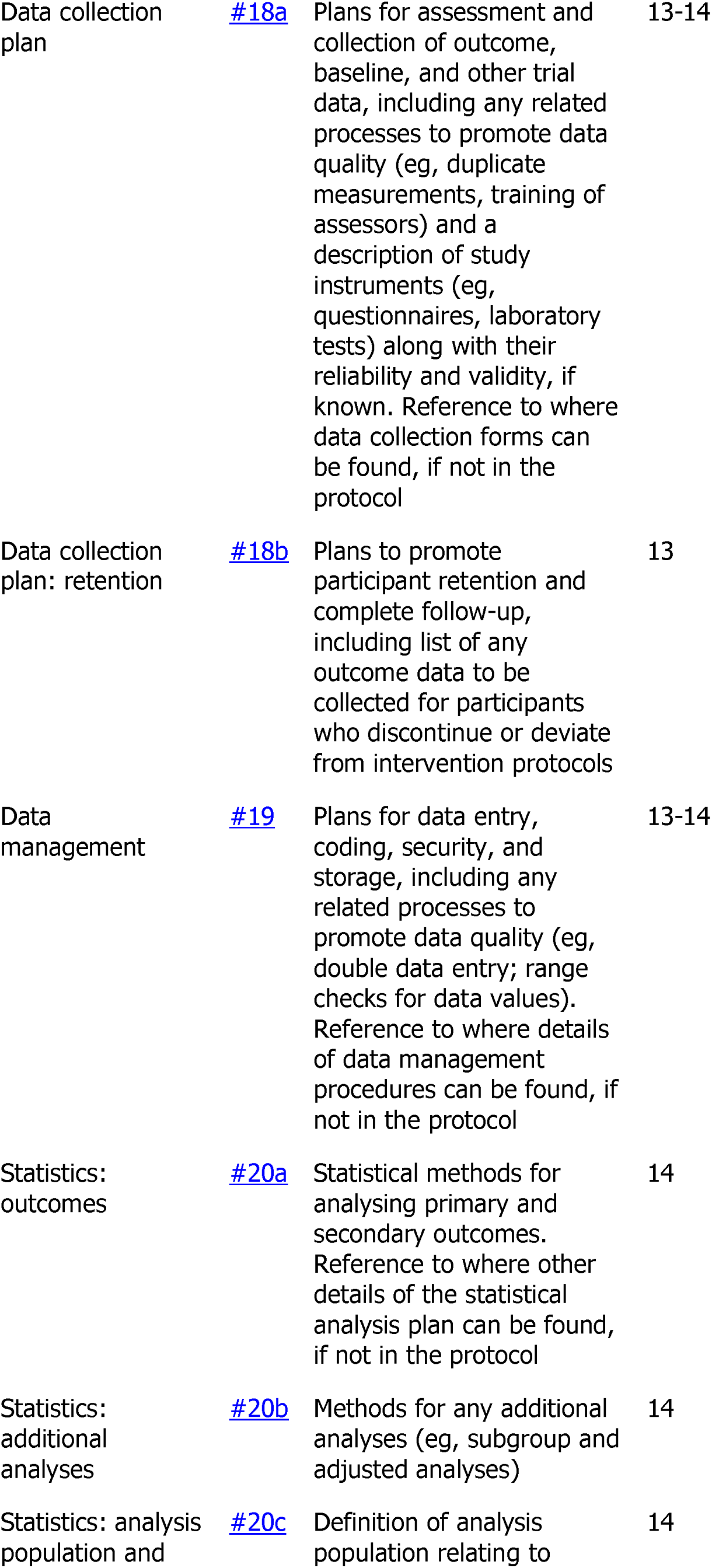
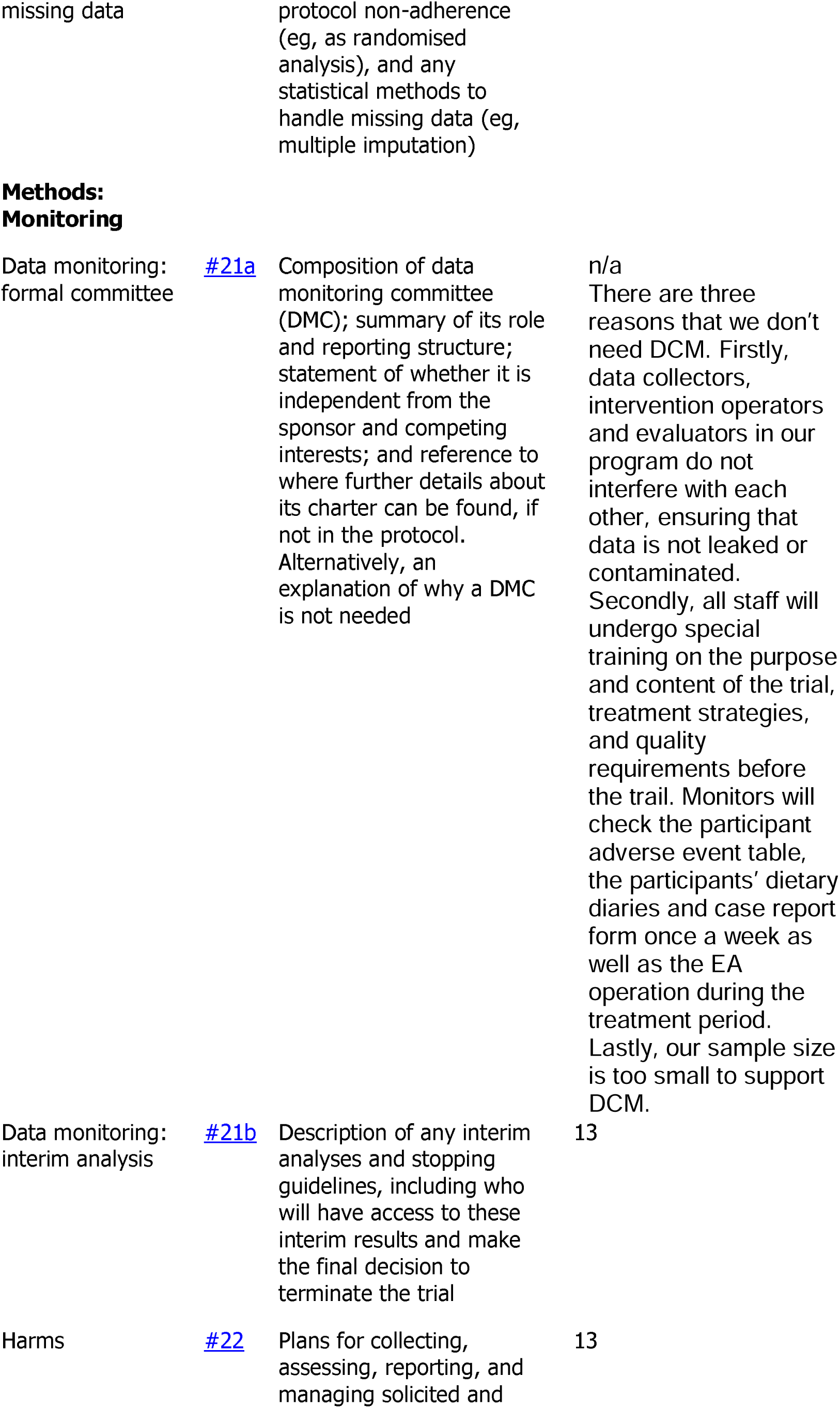
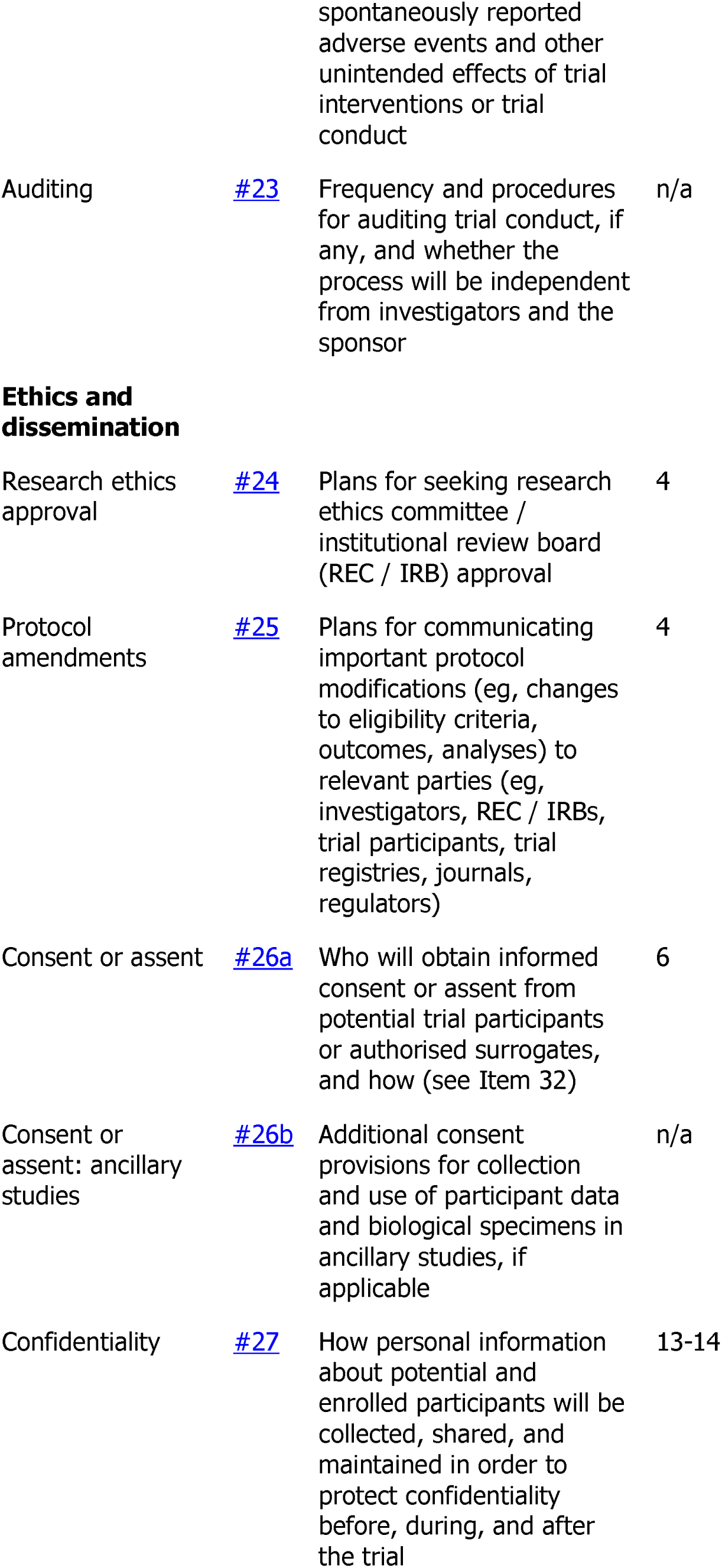
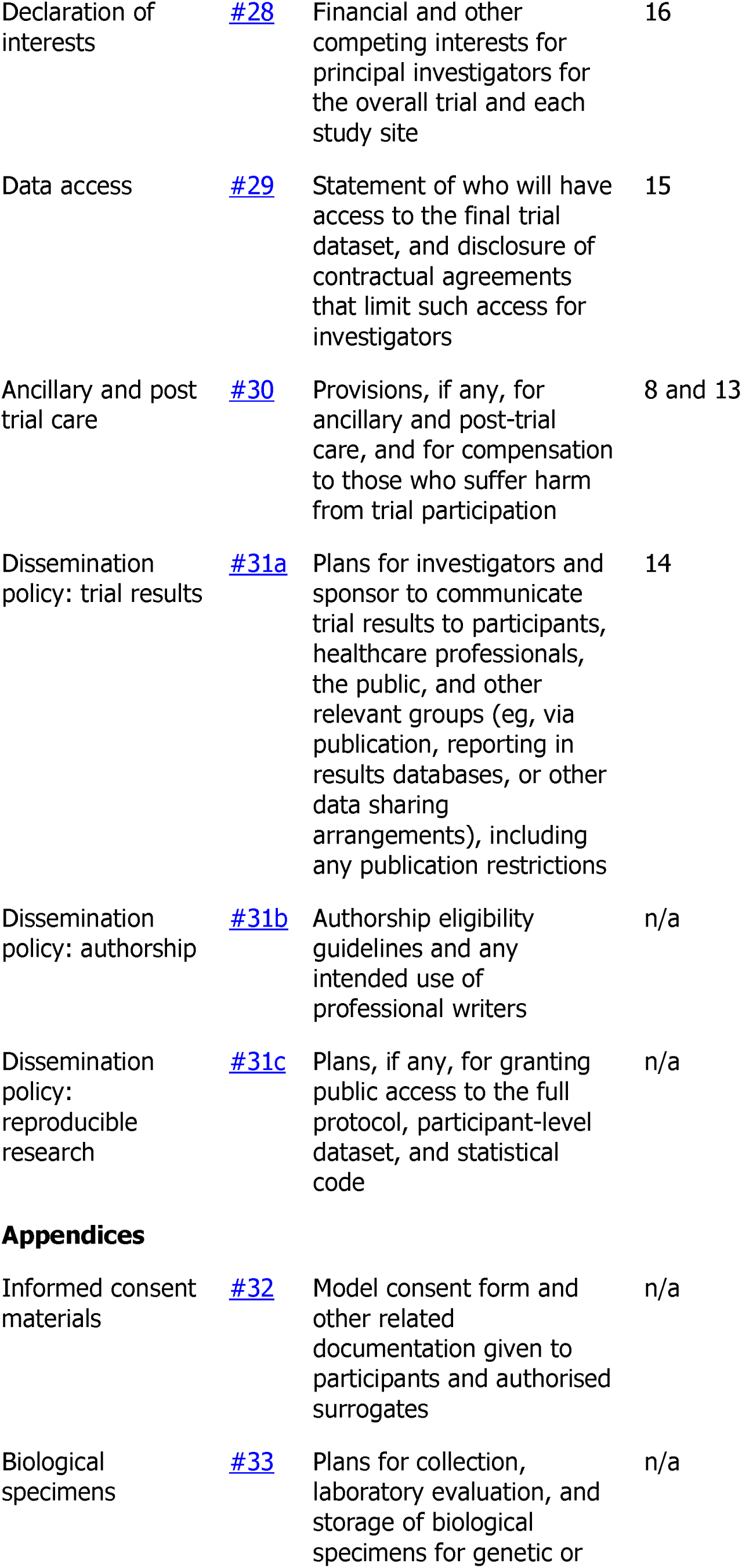
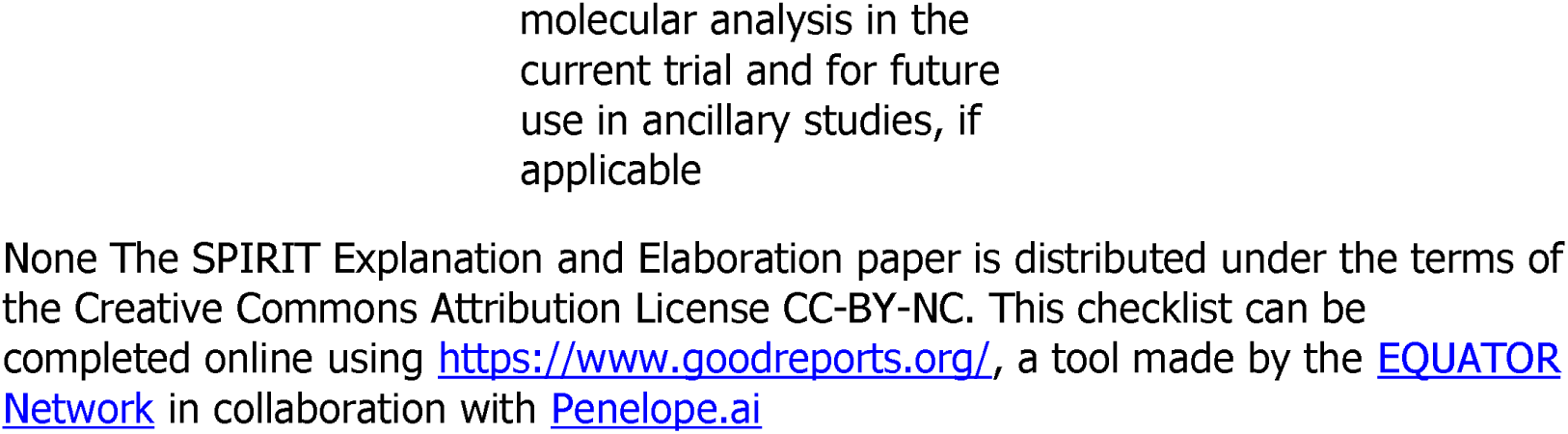
Location of acupoints.

