## Supplementary material for "The central response of electroacupuncture for anxiety in people with obesity based on resting-state functional magnetic resonance imaging: a protocol for a randomized, blinded, sham-controlled trial": https://orcid.org/my-orcid?orcid=0000-0002-7322-0798

Supplementary file 2

| Date: weather： | | | | | |
| --- | --- | --- | --- | --- | --- |
|  | cereals  (g) | meat  （g） | Vegetables and fruits（g） | Dairy products and nuts（g） | Snacks(g) |
| **Breakfast**  time: |  |  |  |  |  |
| **Lunch**  time: |  |  |  |  |  |
| **Dinner**  time: |  |  |  |  |  |
| others | Sleep quality (★/★★/★★★)  Defecation (Yes/NO)  Water intake（1500~1700ml）(Yes/No)  Daily steps (6000) (Yes/No)  Whether to eat after 8:30 pm? (Yes/No) | | | | |
